# Pylorus resection versus pylorus preservation in pancreatoduodenectomy (PyloResPes): study protocol and statistical analysis plan for a German multicentre, single-blind, surgical, registry-based randomised controlled trial

**DOI:** 10.1101/2021.08.05.21260575

**Authors:** Bernhard W. Renz, Christine Adrion, Carsten Klinger, Matthias Ilmer, Jan G. D’Haese, Heinz-J. Buhr, Ulrich Mansmann, Jens Werner, PyloResPres study group

## Abstract

**Introduction:** Partial pancreatoduodenectomy (PD) is the treatment of choice for various benign and malignant tumours of the pancreatic head or the periampullary region. For reconstruction of the gastrointestinal passage, two stomach preserving PD variants exist: pylorus preservation (ppPD), or pylorus resection (prPD) with preservation of the stomach. In pancreatic surgery, delayed gastric emptying (DGE) remains a serious complication after PD with an incidence varying between 4.5% and 45%, potentially delaying hospital discharge or further treatment, e.g. adjuvant chemotherapy. Evidence is lacking to assess which variant of PD entails fewer postoperative DGE.

**Methods and analysis:** The protocol of a large-scale, multicentre, pragmatic, two-arm parallel-group, registry-based randomised controlled trial (rRCT) using a two-stage group-sequential design is presented. This patient-blind rRCT aims to demonstrate the superiority of prPD over ppPD with respect to the overall incidence of DGE within 30 days after index surgery in a German real-world setting. A total of 984 adults undergoing elective PD for any indication will be randomised in a 1:1 ratio. Patients will be recruited at about 30 hospitals being members of the StuDoQ|Pancreas registry established by the German Society of General and Visceral Surgery (DGAV). The postoperative follow-up for each patient will be 30 days. The primary analysis will follow an intention-to-treat approach and applies a binary logistic random intercepts model. Secondary perioperative outcomes include overall severe morbidity (Clavien-Dindo-classification), blood loss, 30-day all-cause mortality, postoperative hospital stay, and operation time. Complication rates and adverse events will be closely monitored.

**Ethics and dissemination:** This protocol was approved by the leading ethics committee of the Medical Faculty of the Ludwig-Maximilians-Universität, Munich (reference no. 19-221). The results will be published in a peer-reviewed journal and presented at international conferences. Study findings will also be disseminated via the website (http://www.dgav.de/studoq/pylorespres/).

**Trial registration number:** DRKS-ID: DRKS00018842 (https://www.drks.de/drks_web/navigate.do?navigationId=trial.HTML&TRIAL_ID=DRKS00018842, prospectively registered on 24-Oct-2019)

**ARTICLE SUMMARY:** **Strengths and limitations of this study**

▪ This will be the first large-scale, surgical registry-based randomised controlled trial (rRCT) in Germany.
▪ Patients will be blinded to the allocated and actually performed treatment.
▪ The active control intervention will be pylorus preserving pancreatoduodenectomy (PD), currently the surgical standard treatment of tumours of the pancreatic head and periampullary region.
▪ Synthesizing evidences from RCTs for this comparative effectiveness research question (preserving vs. resecting PD) is inconclusive with respect to delayed gastric emptying (DGE), a common post-operative complication.
▪ The rRCT approach combines the advantages of a prospective (adaptive two-stage) 2-arm parallel-group RCT and the German multicentre StuDoQ|Pancreas registry as platform for patient enrolment, allocation and data collection, with the aim of improving the evidence-based surgical techniques for PD.

## INTRODUCTION

For resection of malignant or benign lesions of the pancreatic head and the periampullary region, partial pancreatoduodenectomy (PD) is commonly performed.^1,2^ Due to the functional impairment of the physiological propulsive action of the stomach and especially the pylorus, delayed gastric emptying (DGE),^3,4^ with an incidence varying from 4.5% to 45%,^5^ remains a common serious complication after PD. The mechanisms of DGE after PD are multifactorial and still remain unsolved. The left pylorus with its physiological closing function could be a cause. Although not being life-threatening, DGE causes impairment of oral intake often resulting in nausea and vomiting, and is therefore associated with an extended hospital length of stay (LOS), higher healthcare costs and a delay of a potential postoperative adjuvant therapy for tumour patients which may have a negative impact on survival.^6^

There are two gastric resection strategies to perform PD: resection (prPD) or preservation of the pylorus (ppPD). Intriguingly, the benefit of pylorus preservation or pylorus resection in PD with regard to the risk of DGE is still controversially discussed.^5,7,8^ A recent meta-analysis of eight non-randomised studies revealed superiority for prPD compared to ppPD.^5^ However, the subgroup of only three RCTs showed no difference between both procedures regarding DGE. Particularly in the German prospective single-centre PROPP trial, prPD did not reduce the incidence or severity of DGE, but rather tended to have a higher DGE rate.^8^ On the other hand, according to a recent network meta-analysis of randomised trials, prPD seems to be associated with lower rates of DGE.^9^ In line with these inconclusive results, the identified RCTs comparing ppPD and prPD have several limitations arising from clinical and methodological heterogeneity. So far, no high-quality multicentre patient-blinded RCTs exist aiming to prove or disprove the superiority of prPD compared to ppPD for reducing DGE or other major patient-relevant complications.

To fill this evidence gap and to generate clinical knowledge on the safety aspects and performance of different reconstruction techniques in PD from a large-scale multicentre RCT, the registry-based *PyloResPres* trial was initiated. This project is funded by the German Research Foundation (DFG grant number WE 2008/9-0). The German Society for General and Visceral Surgery (DGAV) founded the national pancreatic surgery registry *StuDoQ*|*Pancreas* as a valid platform for risk-adjusted quality assessment, collaborative quality improvement, and outcomes research in pancreatic surgery.^10–13^ This multicentre registry represents a major proportion of the PD patients in Germany at medium- and high-volume centres. Embedding the *PyloResPres* trial in this national registry cohort and employing the already established und implemented electronic case report form (eCRF) is highly efficient and cost-effective in contrast to a conventional RCT.^14^ We present the rationale and design of the *PyloResPres* trial to investigate whether pylorus resection is advantageous over pylorus preservation in reducing the DGE risk and regarding further postoperative patient-relevant outcomes after PD.

## METHODS AND ANALYSIS

This protocol is presented in accordance with the SPIRIT Statement^15^ and also considers the protocol-related aspects of the CONSORT extension for routinely collected data, adaptive designs, and non-pharmacological treatment interventions.^16–18^ The trial will be carried out in accordance with Good Clinical Practice and applicable German legislation.

### Aim of this study

The primary objective of the trial is to determine whether the incidence of DGE within 30 days after index surgery in patients undergoing elective PD is lower with resection of the pylorus (pr) compared to its preservation (pp).

### Study design and setting

PyloResPres is an investigator-initiated, large-scale, multicentre, 2-arm parallel-group, patient-blind, adaptive (two-stage group-sequential), randomised controlled trial conducted in hospitals throughout Germany. This surgical trial is designed as a registry-based RCT (rRCT) to imbed the study population in the spectrum of a real-world PD registry cohort. Its design is also highly pragmatic in the sense of not giving technical instructions on how the pancreatic head resection or the reconstruction should be performed, i.e. limiting deviations from standard of care.

### Patient population and eligibility criteria

This registry-based RCT allows recruiting patients from an unselected target population treated in hospitals of all care levels (basic, regular, and maximum) including specialized high-volume centres. All patients scheduled for an elective PD who are eligible and consented for being registered in StuDoQ|Pancreas will be consecutively assessed for eligibility and informed about the trial details during a pre-treatment visit or on the day of admission to the hospital’s surgical department. The following eligibility criteria were defined for the trial:

#### Inclusion criteria

▪ Age ≥18 years;
▪ planned partial pancreatoduodenectomy for any indication (e.g. resectable benign or malign tumour of the pancreatic head or the periampullary region);
▪ Written informed consent of the patient or legal representative.

#### Exclusion criteria

▪ Age <18 years;
▪ Patients participating in another interventional trial interfering with the surgical intervention or the outcomes of this trial;
▪ Patients with expected lack of compliance and/or irreconcilable language barriers.

By defining less stringent inclusion criteria, almost the whole spectrum of diseases in the pancreatic head region is of interest. The trial population is covered by the multicentre DGAV StuDoQ|Pancreas registry which provides representative data on pancreatic surgery in academic and non-academic hospitals with a medium to high institutional and surgeon caseload, while capturing about 20% of all pancreatic resections performed annually in Germany.^10^ The registry is recognized by surgeons as a valuable tool to assess institutional quality and effectiveness of pancreatic surgery and surgical interventions and has received significant acceptance.

### Site selection

All institutions participating in the StuDoQ|Pancreas registry (currently, about 70 pancreatic surgery centres) are required to meet certain quality standards and are basically deemed eligible.^10,11^ These sites are centres for pancreatic surgery at academic and non-academic community hospitals with a medium to high caseload, at least fulfilling the criteria to participate in the StuDoQ|Pancreas registry.^19^ Ultimately, about 30 registry sites with an annual caseload of about 900 PDs were recruited from the set of eligible hospitals according to their letter-of-intent written during the project application, and further expansion is to be expected if necessary (**Supplemental File 1**).

### Randomisation, allocation concealment and masking

Patients fulfilling the eligibility criteria will be randomly assigned in a ratio of 1:1 to receive one of the two intestinal reconstruction techniques after PD. The randomisation technique is based on permuted balanced blocks and considers stratification by study site. The randomisation list was created by an independent person at the IBE who is not involved into the trial and is concealed from study investigators, other study personnel (e.g. study nurses), data managers and the responsible trial statistician. Neither the investigators nor other trial staffs have access to the randomisation list. Randomisation will be undertaken via the secure, web-based eCRF system REDCap® (Research Electronic Data Capture; Version 9.5.4, Vanderbilt University, Nashville, Tenn.) hosted at the IBE of the LMU Munich (https://random.ibe.med.uni-muenchen.de/redcap/). Depending on local organisational circumstances, randomisation takes place up to one day before the index surgery, or intraoperatively (prior to the step under investigation, i.e. after exploration and mobilisation of the duodenum and the pancreatic head when preservation of the pylorus seems technically and oncologically feasible), after assessing a patient for eligibility and after entering his/her screening data into the official StuDoQ|Pancreas eCRF (i.e., creating a new patient identifier). The randomising clinician or study nurse at the study site will log into the REDCap system that will randomise an eligible patient to the ppPD or prPD group, and will notify the operating surgeon about the intervention allocation for the patient. To verify the actually performed surgical procedure compared to the allocated intended one, the correct randomisation (intended and actual allocation) will be assessed by the histopathological report written by an independent pathologist unaware of the allocated intervention. This report will be reviewed by the monitor for quality control.

By nature of the trial design and the surgical intervention, it is not possible to blind the investigators and study personnel at the hospital. Patients will be blinded to the intervention until discharge and assessment of the primary and secondary endpoints. No attempts will be made to blind the trial statisticians. However, they will not have access to the unblinded eCRF and database during the study and will perform analyses according to a predefined statistical analysis plan (SAP) which will be finalised prior to database closure at latest. Although investigators/healthcare providers, outcome assessors, data managers and data analysts (affiliated to the DGAV and IBE, respectively) will not be blinded, we do not expect a high risk of ascertainment bias to influence the treatment effect for objective outcomes including the primary outcome.^20^

### Surgical interventions

This trial compares two established and routinely performed surgical approaches, with prPD being the experimental procedure and ppPD the control procedure. The board certified surgeon participating in the trial must have the same level of expertise in both techniques to avoid a significant bias due to learning curves.^21^

In both treatment groups, the resection of the duodenum with the head of the pancreas, the first 15−30cm of the jejunum, the common bile duct, the gallbladder, and in oncological cases the locoregional and central lymph node dissection, will be performed according to the local standards. In contrast, the transection of the proximal end of the gastrointestinal tract has to be performed according to the allocated treatment. Furthermore, a pancreatojejunostomy or pancreatogastrostomy and a hepaticojejunostomy will be carried out to connect the pancreas and the bile duct with the intestinal tract in accordance to the standardized approach at the respective study site.

**Figure 1** displays the possibilities of gastrointestinal reconstruction techniques which can be performed in both investigated PD strategies.

**Figure 1.**
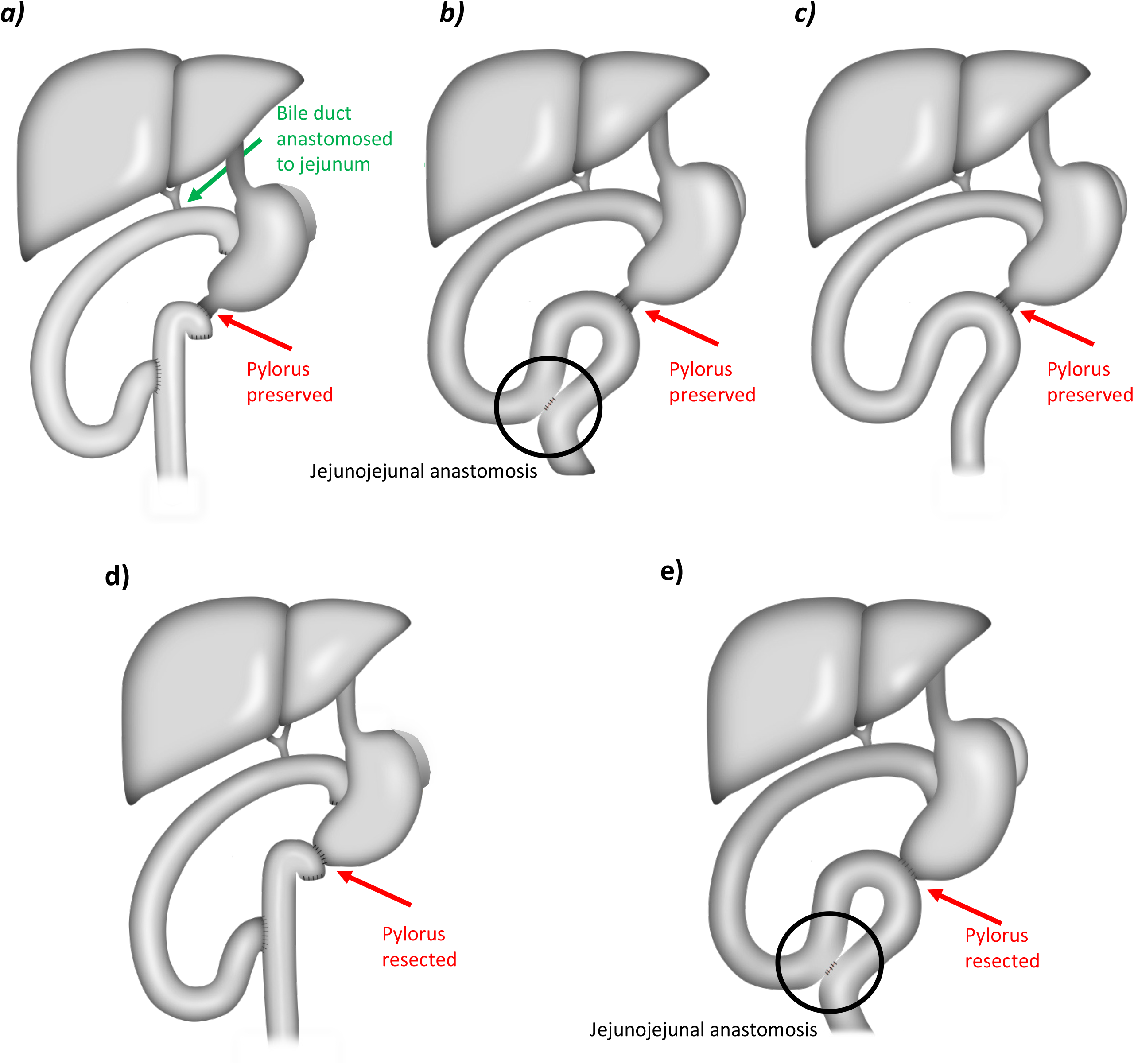
Schematic presentation of possible surgical PD procedures. *Upper panel*: Possible reconstruction methods after ppPD as Roux-en-Y reconstruction **a)**, or as single-loop omega-shaped reconstruction with **b)**, or without **c)** an additional side-to-side jejunojejunal anastomosis. *Lower panel*: Possible reconstruction methods after prPD as Roux-en-Y reconstruction **d)**, or as single-loop omega-shaped reconstruction with an additional side-to-side jejunojejunal anastomosis **e)**. The red arrows indicate pylorus preservation, and pylorus resection, respectively. ppPD = Pylorus preservation in pancreatoduodenectomy; prPD = Pylorus resection in pancreatoduodenectomy.

#### Control intervention: Pylorus preserving partial pancreatoduodenectomy (ppPD)

The duodenum will be divided (e.g. with a linear stapling device) at least 1cm distal to the pylorus, preserving the gastric vessels along the lesser and the greater curvature. After performing the pancreatic anastomosis (pancreatojejunostomy or pancreatogastrostomy) and the hepaticojejunostomy, the gastrointestinal passage can be reconstructed through a standard Roux-en-Y reconstruction or as a single-loop omega-shaped reconstruction with or without an additional side-to-side jejunojejunal anastomosis (**Figure 1a–c**). Regarding the duodenojejunostomy, it is left up to the standard at the respective surgical department whether continuous or interrupted sutures in a single layer or two layer technique are employed, or whether the duodenal staple line is included or excluded from the anastomosis. The route of reconstruction must be documented in the StuDoQ|Pancreas registry eCRF. The ppPD is well-chosen as the reference treatment as it is regarded as the standard treatment for resectable tumours of the pancreatic head and the periampullary region.^22^

#### Experimental intervention: Pylorus resecting partial pancreatoduodenectomy (prPD)

In the prPD group, the stomach will be divided within 5cm proximal to the pyloric ring (e.g. with a linear stapling device) with complete preservation of the gastric vessels along both curvatures to maintain perfusion of the distal stomach via the gastroepiploic vessels and the left gastric artery, respectively. As in the ppPD group, the pancreatic and bile duct anastomosis will be performed. Similar to the control group, reconstruction can be established through a standard Roux-en-Y reconstruction or as a single-loop omega-shaped reconstruction, but in contrast to the ppPD group, an additional side-to-side jejunojejunal anastomosis has to be performed (**Figure 1d and e**). Similar to the ppD group, the technique of the duodenojejunostomy will be left up to the preferences of the participating surgery department. The route of reconstruction must also be documented in the eCRF.

The same perioperative and postoperative standard operating procedures are in place for both surgical interventions, and study patients in both groups will be accommodated on the same wards to ensure postoperative care according to the institutional standards.

#### Conversion from ppPD to prPD, adherence to the assigned intervention

Due to intraoperative circumstances and disease severity, not all patients randomised will receive the assigned intervention reflecting clinical reality. This is particularly true in the case of pre-operative randomisation. If a patient is randomised on the day before surgery to receive ppPD, the surgeon will have to confirm that PD is feasible, and preservation of the pylorus is technically possible and oncologically justified. Otherwise, the surgeon will have to convert from ppPD to prPD. Reasons for conversion can arise intraoperatively, e.g., for technical infeasibility or significant bleeding. Affected trial participants will be denoted as ‘crossover patients’. In the case of a pre-operative randomisation, a patient can also turn out to be non-resectable or having a previously unknown metastatic disease, and neither of the two surgical procedures investigated can be undertaken. The reason for all deviations between the intended and actual allocation will be documented in the eCRF. If a pre-operatively randomised patient does not receive the intended treatment or even neither of the treatments, he or she remains in the trial until the end of the follow-up period.^21^

#### Relevant concomitant care during the trial

Due to the pragmatic attitude of this rRCT, no instructions or recommendations on the perioperative management of the patients will be given. Hence, no permitted or prohibited procedures exist. Peri- and postoperative medical treatment including antibiotic medication, prokinetic drugs and somatostatin analogues should be administered according to the respective institutional standards, but must be documented in the registry. Furthermore, endoscopic examination of the duodenojejunostomy or the gastrojejunostomy in the case of DGE can be performed. In both groups, the nasogastric tube can be removed as soon as mechanical ventilation is stopped, but the decision remains up to the surgeon in charge. All patients will be further treated as it is routinely carried out in the respective surgical department.

### Study outcomes and data collection schedule

#### Primary outcome

The primary outcome is the occurrence of DGE grade A, B, or C (mild, moderate, and severe forms) up to 30 days post-surgery. The assessment of DGE is objective and uses the obvious criterion of the presence of a gastric tube as defined by the ISGPS in 2007.^4^ **Table 1** displays the different forms of DGE classified by their clinical impact. All study patients should be classified with result DGE yes vs no (irrespective of the clinical grading). A patient will be positive, if DGE is diagnosed within 30 days after index operation. In both treatment groups, the overall DGE incidence within 30 days after the index operation will be assessed as primary outcome measure. We chose the time period of 30 days to reliably observe all occurring postoperative events of DGE because after this time interval it is very unlikely that reinsertion of the nasogastric tube is causally linked to DGE. Several trials have already used this primary endpoint.^23^

**Table 1.** Delayed gastric emptying (DGE) clinical grading after pancreatic resection according to the International Study Group of Pancreatic Surgery (ISGPS) consensus definition.^4^

| DGE grade | Nasogastric tube required | Unable to tolerate solid oral intake by POD | Vomiting/ gastric distension | Use of prokinetic medication |
| --- | --- | --- | --- | --- |
| A | 4-7 days, or reinsertion > POD 3 | 7 | ± | ± |
| B | 8-14 days, or reinsertion > POD 7 | 14 | + | + |
| C | >14 days, or reinsertion > POD 14 | 21 | + | + |

POD, postoperative day. ± means uncommon/ possible, + means present/ necessary.
The mild, moderate, and severe forms of DGE after pancreatic resection can be classified into grades A, B, and C by their clinical impact.
Reinsertion is documented in the eCRF with time of reinsertion and duration of the treatment.

#### Secondary outcomes

To compare peri- and postoperative outcomes between both groups, the following patient- and surgeon-oriented secondary outcomes will be assessed:

#### Intraoperative outcomes

▪ Operation time [minutes],
▪ Histological parameters
▪ Conversion from ppPD to prPD [yes/no]
▪ Blood loss [ml],

#### Postoperative outcomes

▪ General complications within 30 days after index operation (e.g. wound infection, pulmonary complications, organ failure) scored according to the Clavien-Dindo classification (<3b vs ≥3b);^24,25^
▪ Overall mortality: death from any cause within 30 days after index operation;
▪ Postoperative hospital length of stay (LOS): Number of days from the day after index surgery up to the day of discharge from primary hospital;
▪ Insufficiency of gastro-/ duodenojejunostomy and the jejunojejunal anastomosis [yes/no] within 30 days after index operation;
▪ Pancreatic surgery-specific complications (besides DGE) within 30 days after index operation:
  - Postoperative pancreatic fistula^26^
  - Postpancreatectomy haemorrhage^27^
  - Biliary leak^28^
  - Chyle leak/ lymphatic fistula;^29^
▪ Necessity for re-operations: Number of re-operations within 30 days after index operation;
▪ Hospital re-admissions: subsequent hospital re-admission [yes/no] within 30 days after index operation.

#### Data collection and trial visits

During the screening visit (≥1 days before surgery), eligibility criteria will be assessed. Those patients who refuse written informed consent for the trial but give informed consent for the StuDoQ|Pancreas registry will be documented in the registry only. After providing informed consent to participate in the trial, the patient’s demographic and clinical characteristics, medical history, comorbidities, preoperative diagnostics and laboratory will be assessed and documented. After randomisation, surgical intervention will occur on day 0 with assessment of the operation details, intra-and perioperative parameters as well as adverse events. A standard follow-up care will be performed. Follow-ups are scheduled on day 7, day 14 or the day of discharge (whichever occurs first), and on day 30 after study-related index operation. Postoperative follow-up can occur as clinic visits or by phone with documentation of the primary and secondary endpoints including serious adverse events. **Table 2** summarizes the patient data collection. **Figure 2** displays the patient flow.

**Table 2.**
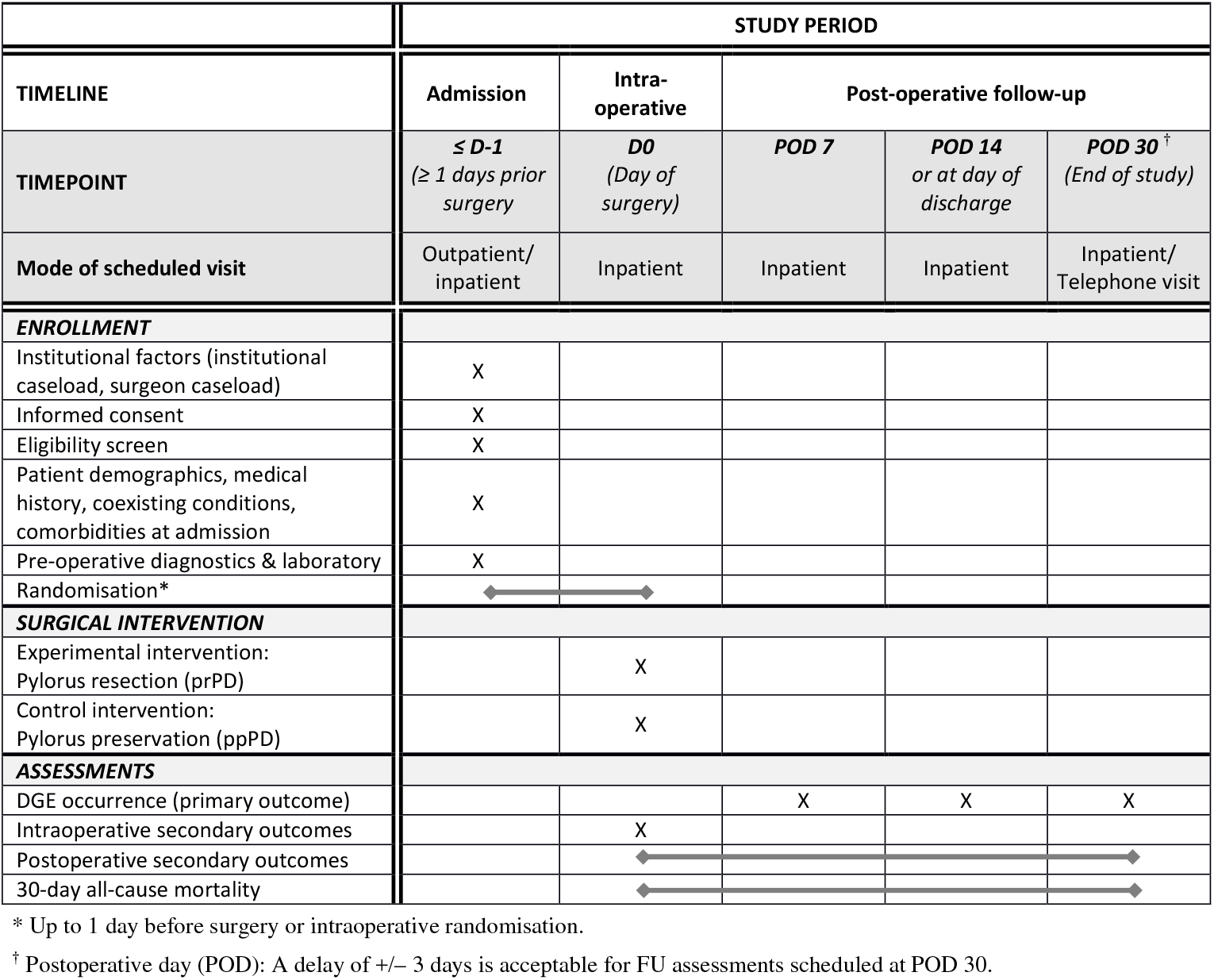
Schedule of enrolment, interventions and assessments in the *PyloResPres* trial.

**Figure 2.**
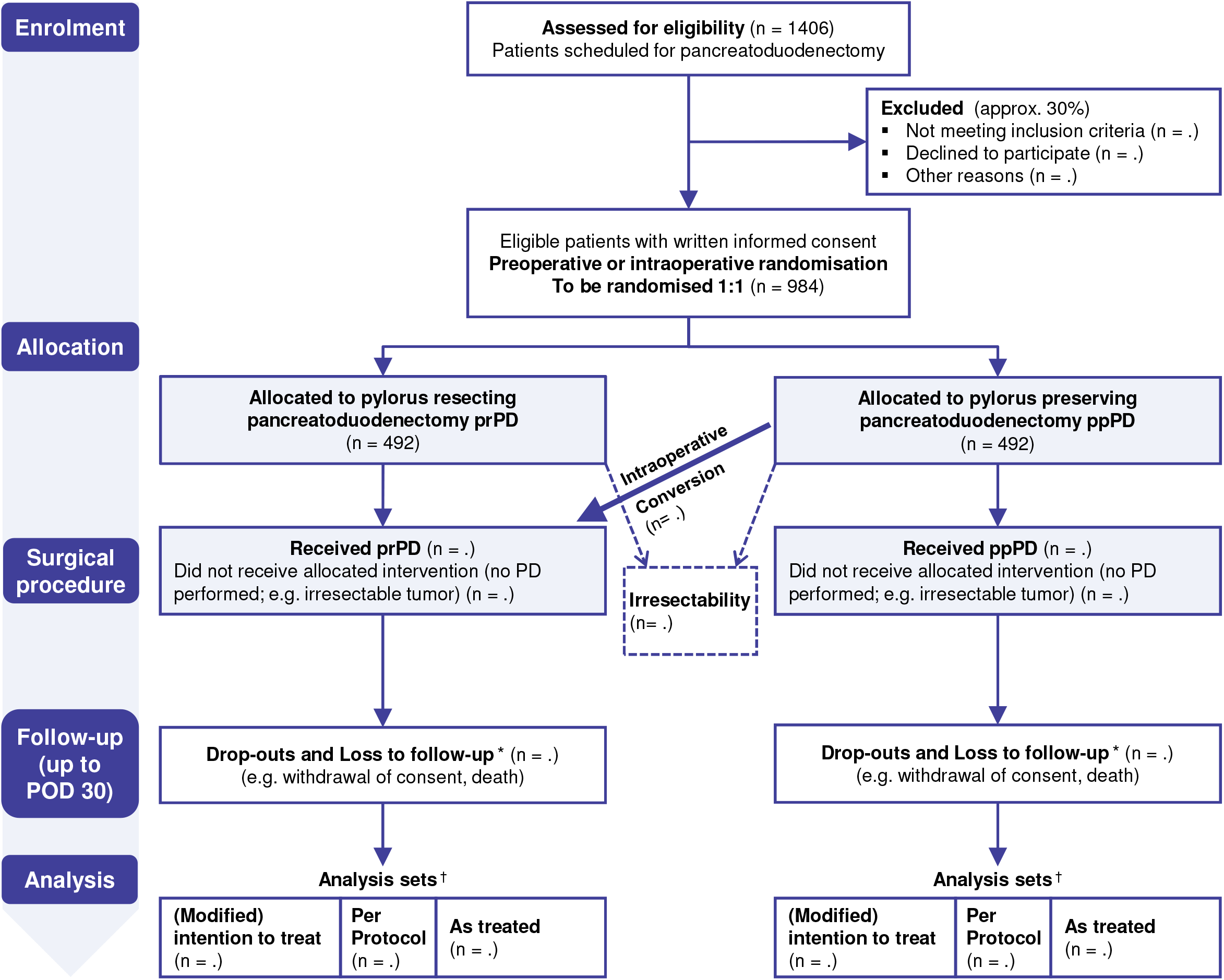
CONSORT flow chart describing the planned trial process (ignoring the planned interim analysis). * Loss to follow-up estimated to be marginal during the follow-up period until POD 30. ^†^ Exclusion from analysis estimated to be marginal during the follow-up period (until POD 30). 2-stage written informed consent (documentation in the StuDoQ|Pancreas registry; enrolment into the trial). CONSORT, Consolidated Standards of Reporting Trials; POD, postoperative day.

Outcome assessors will evaluate the presence of the primary endpoint during clinic visits and from the patient’s medical record and, if indicated, by a telephone call on POD 30.

#### Safety aspects

Complication rates and SAEs will be closely monitored for the assessment of safety. SAEs occurring in the period from the day of randomisation to the end of the postoperative follow-up will be documented.

### Data management, data security and quality control

#### Registry-based reporting and data management

This trial is nested within the StuDoQ|Pancreas registry documenting key parameters related to pancreatic surgeries since September 2013 and uses the same infrastructure. Data acquisition is performed by a browser-based eCRF system which is based on PHP and R software.^10^ All study data will be stored electronically in the StuDoQ|Pancreas registry.

In summary, using the IT-infrastructure of the registry provides the following advantages: (1) Participation of institutions and documentation is already implemented. (2) The eCRF is already established und implemented. (3) Additional trial-specific items can be added at any time. (4) The data management processes (including automated plausibility checks of data entry and query generation) are already established and were extended for trial purposes implementing further consistency checks. (5) Expenses for implementation of eCRFs and data management are minimal. (6) The study population can be reflected within the documented, more general patient population increasing the generalizability of the trial results.

All installations were made in compliance with the GDPR and are continuously monitored by the data protection officers. All study data will be entered by the investigators or designated staff into the StuDoQ eCRF and stored in pseudonymized form. Data management and quality control will be performed using automatic data validation requests/ data entry plausibility checks within the eCRF and on-site monitoring to ensure accuracy and enquire implausible or missing data on a regular basis. Supplementary periodic validation checks performed by the data management team will also encompass data derived by the randomisation tool REDCap and will take place on a regular basis. After web-based data entry into the StuDoQ eCRF, data are automatically checked for completeness and plausibility. Completeness of major quality indicators is mandatory for the inclusion in the annual institutional reports and DGAV certification purposes. Missing or implausible items generate warning messages and queries. Besides, the experience of recruiting centers is documented in the annual quality reports of the registry (**Supplemental File 2**).

A formal protocol of the registry detailing aims and regulations concerning informed consent, data safety and publication issues was established by the DGAV and can be accessed at http://www.dgav.de/studoq/datenschutzkonzept-und-publikationsrichtlinien.html (in German). This set of rules governing the registry therefore applies to this trial as well. These strategies for data protection regulation are based on the guidelines set forth by the Society for Technology, Methods and Infrastructure for Networked Medical Research (TMF).^30^ The approved concept of “decentralised patient lists” of the TMF is applied, which allows re-identification only by the treating participating hospital.

#### Monitoring

As most study centres already participate in the StuDoQ|Pancreas or another organ-specific StuDoQ registry, a high level of routine for data entry and verification can be assumed. As the initiation of the first centres happened closely after the outbreak of the COVID-19 pandemic, sites were initiated by virtual meetings. As part of a risk-based monitoring strategy, all centres will be visited on-site or virtually after enrolment of the first 10-15 patients. Key data will be verified for 100% of patients. Trial sites will then be assessed as being either with or without noticeable findings with respect to the trial protocol, data quality and GCP. Monitoring will be performed by the study centre at the site of the coordinating investigator at LMU Munich. The monitor (affiliated to the sponsor of the trial) reviews the eCRF for completeness and accuracy and instructs site personnel to make any required corrections. During monitoring visits, the monitor will ensure that data documented in the eCRF are in line with underlying source data. Sites without noticeable problems will receive no further on-site monitoring visits. Centres assessed as having noticeable problems will be visited again within four months. If problems persist, visits will be repeated three times per year, and key data will be verified for at least 50% of patients at the respective site. If no more findings occur, monitoring visits will no longer be required.

### Data monitoring committee

An independent data monitoring committee (DMC) has been established comprising of one clinical expert in this field and one biostatistician. The DMC will monitor the trial progress as well as the protocol adherence. All preliminary reports and the interim analysis will be discussed and reviewed for accuracy and conclusiveness. The DMC will make recommendations to the principal investigator to continue or terminate the study on the basis of the results from the pre-planned interim analysis. The board will meet at the beginning of the study and at 6-months intervals thereafter, or as needed.

### Patient and public involvement

No patients were involved in setting the research question or the outcome measures, nor were they involved in developing plans for participant recruitment, or the design and implementation of the study. However, the rationale and objectives of this trial are of major concern for the target patient population and have been a topic for many years in patient organizations such as the German AdP (Arbeitskreis der Pankreatektomierten e.V.).^31^

### Statistical methodology and planned analyses

#### Power considerations and pre-trial sample size calculation

Sample size calculation was performed on the basis of the primary outcome: occurrence of DGE up to POD 30. A recent meta-analysis of RCTs reports a DGE rate of 18% (95% CI [13%; 23.4%]) for prPD, and 21.5% (95% CI [15.6%; 27.1%]) for ppPD patients.^5^ Based on this result and our aim to detect a clinically relevant difference between prPD and ppPD, we chose the following planning scenario: An absolute risk difference of 10% in DGE between both groups is regarded as a minimum clinically important effect, with 25% DGE risk in the ppPD and 15% in the prPD group. We expect that for 15% of the patients in each group the corresponding DGE outcome cannot be assessed (e.g. due to dropout, lost to follow-up or mortality). In this case, the binary outcome will be set to failure within the analysis. Therefore, a failure rate of 0.30 (=0.15+0.15) for the prPD and of 0.40 (=0.25+0.15) for the reference group ppPD was assumed for the initial sample size calculation.

Due to the possible imprecision of the reported rates, a two-stage group sequential design was set up, with one interim analysis after assessment of 50% of the target sample size. The critical values and the test characteristics of this group sequential test design were calculated for a Wang & Tsiatis design with boundary shape parameter Delta=0.23 (values of Delta between 0 and 0.5 give critical values ranging between Pocock and O’Brien & Fleming tests).^32,33^ For a pre-specified significance level of 5% (two-sided) and event rates π_prPD_=0.3 and π_ppPD_=0.4 (odds ratio (OR) of 1.556), the power is 90.0% if both stages consist of two times 246 patients randomised per group, thus resulting in a total number of 984 patients to be assigned to the trial (**Supplemental file 3)**. Assuming that about 70% of eligible patients consent to study participation, about 1406 patients in total have to be screened (**Figure 2**).

#### Efficacy interim analysis and continuous study planning

As soon as the primary endpoint has been assessed for 492 randomised patients in total (50% information fraction), the interim analysis will be conducted to inform potential adaptations, with futility stop based on the pre-defined boundaries. If the study continues to the second stage, sample size adaptation will be applied according to Müller & Schäfer,^34^ while controlling the overall 2-sided 5% significance level.

To compensate for the loss of power due to post-randomisation dropouts (non-resectable/ metastasized patients not being part of the principal analysis) and crossover patients (who did not receive the allocated surgical procedure (prPD instead of ppPD)), which may diminish the observed treatment difference, it might become necessary to increase the sample size accordingly within the scope of the interim analysis aiming to achieve 90% power when analysing the per protocol set as the smallest population.^21^ A certain proportion of crossover patients being unavoidable in the case of pre-operative randomisation was not considered in the initial sample size calculation as no data were available, and the extent of the conversion rate is difficult to be foreseen.

#### Premature closure of the trial

Stopping is statistically based on rules derived from the primary interim result, however, also based on the feasibility of patient accrual. The trial will be stopped if the first-stage p-value resulting from the planned interim analysis lies below 0.0143 for efficacy to ensure the statistical validity of the applied two-stage group sequential design. It will also be stopped prematurely if the interim analysis permits continuation of the trial, but the re-calculated sample size to achieve sufficient statistical power is infeasible or inefficiently high. Thus, this design avoids prolonged recruitment in order to reach statistical power if the goal is unachievable (the interim analysis cannot account for misspecification of the design parameters that may emerge from the interim results) or, if early significance can be claimed.

#### Statistical analysis plan

The primary efficacy analysis will be based on three analysis sets. Patients deemed unresectable during surgery (e.g. total pancreatectomy performed) will not be considered in any of the analysis sets, as the primary aim of this trial is to estimate the intervention effect after a PD.

The modified intention-to-treat (mITT) set (FAS, full analysis set) comprises all patients in the group to which they were randomised (converted patients remain in the intended ppPD group; non-resectable patients not considered). Analysing the mITT set can be interpreted as a treatment policy approach according to the estimands framework.^35^ The per-protocol (PP) set consists of all patients treated per protocol; patients with major protocol violations and crossover patients will be excluded, and no missing data will be imputed (non-resectable patients not considered). In addition, the As-Treated (AT) set will be analysed considering patients according to their actual, rather than randomised (intended) treatment (i.e., converted patients analysed in the prPD group; non-resectable patients not considered). Non-resectability is handled by excluding those patients from all the analyses as they are not part of the targeted population. Other post-randomisation events, like resection of adjacent organs, will be ignored.

##### Confirmatory analysis of the primary endpoint

The null hypothesis for the primary binary endpoint assumes that the 30-day DGE rate is equal in the ppPD and prPD group. To account for the factor site used for stratifying the randomisation, the principal model will include random intercepts for study site. To compare DGE rates between both groups in the final analysis, a logistic random intercept model will be applied with treatment as fixed effect, adjusting for the surgeon caseload (≥20 vs <20 procedures per year). The effect of the intervention will be presented as the OR of DGE for prPD vs ppPD, together with its 95% CI. In addition, crude proportions (DGE incidence) by treatment arm will be reported with an unadjusted OR and 95% CI, and a χ^2^ test p-value. If not revised in the SAP, missing values will be replaced by means of the ICA-r method proposed by Higgins *et al*. (Imputed Case Analysis incorporating available reasons for missing data).^36^ The pre-planned interim analysis will be done in accordance with the ITT principle in the same way as the final analysis.

##### Analyses performed for secondary outcome measures and safety

Concerning secondary endpoints on clinical effectiveness, exploratory analyses will be performed reporting appropriate summary measures as well as descriptive two-sided p-values. Random intercept models will be applied: logistic regression for binary perioperative outcomes (e.g. 30-day all-cause mortality); linear regression for log-transformed operation time and other continuous secondary outcomes. These models will also include caseload as adjusting factor and as additional fixed effect. The safety analysis includes calculation and comparison of frequencies and incidences of all complications and SAEs in the prPD vs. ppPD group. These safety data will be tabulated and descriptively summarized for each time point.

##### Exploratory subgroup and registry-specific analyses

Exploratory subgroup analyses (subgroups e.g., tumour patients, extension of resection, the patient’s BMI, age) will be performed to investigate the heterogeneity of the treatment effect across both surgical groups within the target population. Registry-specific analyses aim to describe the trial population within the relevant StuDoQ|Pancreas registry population to assess selection processes at different levels.^37^ Further, a subgroup analysis will be performed for DGE grade A, and grade B or C combined (considered being clinically relevant DGE). In a supplementary analysis, the impact of treatment in hospitals of different care levels (basic, regular, and maximum) regarding DGE incidence will be assessed and compared between both surgical groups.

Statistical analyses will be performed using the software package R version 4.1.0 or higher.^38^ A full SAP will be written ahead of the final database lock and commencement of analysis.

## CONCLUSION

As the first surgical registry-based RCT in Germany, PyloResPres is the largest multicentre trial to date to evaluate the impact of two competing pancreatic head resection strategies (pylorus-reserving vs. - resecting PD) on DGE in patients with any indication for surgery of the pancreatic head or the periampullary region and eligible for both techniques. It is embedded in the national StuDoQ|Pancreas registry which is used as a platform for electronic case records and 30-day standard-of-care postoperative follow-up by minimizing visits outside normal practice and thus limiting additional workload. This novel trial concept enables inclusion of many eligible patients in a relatively short time with limited additional effort and expenses, and facilitates generalizability. Since well-defined baseline characteristics and risk factors of non-enrolled patients are automatically documented in the registry-based eCRF as well, it will be possible to project the results of this rRCT on the entire registry population to assess selection bias.

## TRIAL STATUS

At the time of first manuscript submission, research ethics approval has been obtained for the trial. The study was presented at a meeting with all participating centres during the annual DGAV conference in Wiesbaden, Germany, in October 2019. Data collection with enrolment of the first patient commenced on 25 November 2019 at the site of the coordinating investigator. Accrual of 492 patients randomised to perform the pre-planned interim analysis is expected by the end of the year 2021. Enrolment can be completed by spring 2023, but may change depending on an adaptation decision made after the interim analysis.

## ETHICS AND DISSEMINATION

### Ethical considerations

This study is being conducted in accordance with ethical principles that have their origin in the Declaration of Helsinki and are consistent with Good Clinical Practice (GCP). Enrolment of patients at the participating hospitals did not start until the written and unrestricted positive vote of the local ethics committee (EC) was obtained. The protocol is based on the underlying project application which received previous independent peer review as part of the grant funding process. Together with the patient information sheets and consent forms the protocol was first approved by the primary EC of the Medical Faculty of the Ludwig-Maximilians-Universität, Munich, Germany on 14 August 2019 (approval number 19-221). Amendments to the protocol will be submitted to the EC for review. Individual written informed consent including consent to registry-based data collection will be obtained from all eligible patients in the trial. Consent forms for the trial include consent for publication of results in peer-reviewed journals. Relevant data protection rules for all analysed data will be enforced.

### Dissemination

Following trial completion, data will be available for future secondary-use by participating centres of the registry. The main results will be reported according to relevant guidelines such as the CONSORT extension for trials using routinely collected data and the IDEAL framework and recommendations for the evaluation of surgical interventions in RCTs (Stage 3),^16,39,40^ and will be published in international peer-reviewed scientific journals in this field. Authors and collaborators will be involved in reviewing drafts of the manuscripts arising from this trial. Additionally, these results will be considered in German treatment guidelines. Relevant information about the trial results will be disseminated to patient groups (e.g., the German ‘Arbeitskreis der Pankreatektomierten e.V.’) and the trial website (http://www.dgav.de/studoq/pylorespres.html).

## Supporting information

PyloResPres__SupplementalFile_1_StudyGroup

PyloResPres__SupplementalFile_2_StuDoQ Registry

PyloResPres__SupplementalFile_3_StatisticalMethods

SPIRIT Checklist

## Data Availability

All data of the StuDoQ|Pancreas registry are stored in pseudonymized form, and any re-identification can only be done by the treating, participating hospital. Individual participant data cannot be shared publicly because participants did not explicitly consent to the sharing of their data as per the European Union's General Data Protection Regulation (GDPR) and the corresponding German privacy laws. De-identified participant data and a data dictionary defining each field in the dataset will be made available to researchers upon formal request and receipt of a signed data sharing agreement in accordance with the data sharing policies of the LMU Munich, Germany. Data will be available through the Research Ethics Board of the LMU Munich for researchers who meet the criteria for access to confidential data and after approval of a methodologically sound research proposal. Requests for data access can be directed to and the Data Use and Access Committee (DUAC) of the Medical Faculty of the University of Munich.

## List of Abbreviations

(S)AE(s): (serious) adverse event(s)
(e)CRF: (electronic) Case report form
DGAV: German Society for General and Visceral Surgery (Deutsche Gesellschaft für Allgemeinund Viszeralchirurgie)
DGE: Delayed gastric emptying
DRKS: German Clinical Trials Register
EC: Ethics committee
EDC: Electronic data capture
FAS: Full analysis set
FPF(L)V: First patient first (last) visit
GDPR: General Data Protection Regulation
ICH-GCP: International conference for harmonisation (of technical requirements for pharmaceuticals for human use) − good clinical practice guideline
ICU: Intensive care unit
ISGPS: International Study Group of Pancreatic Surgery
ITT: Intention to treat
LPF(L)V: Last patient first (last) visit
PD: Partial pancreatoduodenectomy
POD: Postoperative day
PP: per protocol
ppPD: Pylorus preservation in pancreatoduodenectomy
prPD: Pylorus resection in pancreatoduodenectomy
QoL: Quality of life
REDCap: Research Electronic Data Capture
rRCT: registry-based randomised clinical trial
SPIRIT: Standard Protocol Items: Recommendations for Interventional Trials
StuDoQ: Study, Documentation and Quality centre (Studien-, Dokumentations-und Qualitätszentrum) of the DGAV

## DECLARATIONS

### Patient consent for publication

Not required.

### Authors’ contributions

BR and JW did the clinical conceptual planning. UM designed the trial and has oversight for the statistical analyses; UM and CA conceived the statistical analysis plan. JW, BR, UM and CA contributed to the preceding research proposal leading to the funding of the trial and were involved in the protocol development and trial setup. JW led the grant application and, as coordinating investigator and sponsor delegated person, has oversight for the trial. HB is responsible for the registry infrastructure. BR is the study project manager, responsible for the study’s quality assessment, and he and JW are in charge of the overall trial management. As information scientist, CK is operating the registry and responsible for data management, data verification and data quality assurance of the official study database being part of the StuDoQ|Pancreas registry.

CA and BR drafted the manuscript (shared first authorship).

MI and JD’H revised the manuscript critically for important intellectual content.

The corresponding author attests that all listed authors meet authorship criteria and that no others meeting the criteria have been omitted. All authors have critically read, contributed with inputs and revisions, and approved the final manuscript.

## Acknowledgements

We are very grateful to the clinical staff of the participating sites involved in the set-up, recruitment and data acquisition (see online **supplementary file 1**).

Notably, we would like to acknowledge the patients for their willingness to participate in this research. We are grateful to Thorsten Hahn and Cristina Burkert, both study assistants at the KCS (Koordinationszentrum Chirurgische Studien) at the site of the coordinating investigator, for their effort and project management activities. We also thank Arthur Gil (IBE, LMU Munich) for implementing the REDCap-based randomisation tool. Further, we would like to thank the following members of the independent Data Monitoring Committee: Prof Marc Besselink, MD (professor of pancreatic and hepatobiliary surgery, Department of Surgery, Amsterdam UMC, University of Amsterdam, Netherlands), and Thomas Bruckner, PhD (biometrician, Institute of Medical Biometry and Informatics, University of Heidelberg, Germany).

## Ethics approval

Ethical approval for the protocol has been granted from each of the participating medical facilities involved in this trial prior to patient recruitment (leading ethics committee of the Medical Faculty of the Ludwig-Maximilians-Universität, Munich, Germany; reference no. 19-221). This paper presents the protocol version 3.0, 22.09.2020.

## SUPPLEMENTARY DATA

**Supplementary file 1. Organizational structure with full list of *PyloResPres* study group members**

**Supplementary file 2. StuDoQ**|**Pancreas registry - further information regarding data acquisition**

**Supplementary file 3. Statistical Methods – further details**

