## Supplementary material for "Pylorus resection versus pylorus preservation in pancreatoduodenectomy (PyloResPes): study protocol and statistical analysis plan for a German multicentre, single-blind, surgical, registry-based randomised controlled trial": PyloResPres__SupplementalFile_1_StudyGroup

### SUPPLEMENTARY MATERIALS

### Organizational structure

#### Trial oversight and management

The sponsor-delegated person and principal investigator is the clinical lead of the Department for General, Visceral and Transplantation Surgery, Ludwig-Maximilians-Universität, Munich. With its trial management office for surgical trials (KCS, Koordinationszentrum Chirurgische Studien), this department is the coordinating centre of this trial, accompanies the study and, during regular meetings, may deal with any relevant project issues.

#### Study Group

##### Site selection

Study sites are centres for pancreatic surgery at academic and non-academic community hospitals with a medium to high caseload, at least fulfilling the criteria to participate in the StuDoQ|Pancreas registry, or already being certified for pancreatic surgery by the German Society of General and Visceral Surgery (DGAV), or participating in this trial in order to meet certification standards as accredited by the DGAV in Germany. Participation of hospitals in the StuDoQ registry is voluntary. Since 2017, participation in the registry has become mandatory for institutions that aim to meet certification standards of the DGAV.

##### Investigators and participating hospital facilities

At the time of this manuscript submission, 27 registry sites throughout Germany gave consent to participate in the trial and were initiated for patient recruitment. Further trial centres will be recruited in the near future. The principal investigators of the participating centres are listed in the following.

***Munich:***
**Hospital of the Ludwig-Maximilians-Universität**
Department for General, Visceral and Transplantation Surgery
Marchioninistr. 15, 81377 Munich, Germany
Prof Jens Werner, MD (**Coordinating investigator, Trial Sponsor/ Sponsor Delegated Person)**

*Erlangen:*
**University Hospital of Friedrich Alexander University Erlangen-Nürnberg (FAU)**
Department of General and Visceral Surgery
Krankenhausstr. 12, 91054 Erlangen, Germany
Maximilian Brunner, MD

*Würzburg:*
**University Hospital of Würzburg**
Department of General, Visceral, Transplant, Vascular and Pediatric Surgery
Oberdürrbacher Str. 6, 97080 Würzburg, Germany
Stefan Löb, MD

*Fulda:*
**Klinikum Fulda**
Department of General and Visceral Surgery
Universitätsmedizin Marburg – Campus Fulda
Pacelliallee 4, 36043 Fulda, Germany
PD Achim Hellinger, MD

*Memmingen:*
**Klinikum Memmingen**
Department of General, Visceral and Thorax Surgery
Bismarckstr. 23, 87700 Memmingen, Germany
Daniel Krampulz, MD

*Rosenheim:*
**RoMed Klinikum Rosenheim**
Department of General, Visceral and Thorax Surgery
Pettenkoferstr. 10, 83022 Rosenheim, Germany
Prof Katja Ott, MD

*Essen:*
**Alfried Krupp Krankenhaus Rüttenscheid**
Department of General and Visceral Surgery
Alfried-Krupp-Straße 21, 45131 Essen, Germany
Prof Marco Niedergethmann, MD

*Köln:*
**St. Vinzenz-Hospital Köln –** Lehrkrankenhaus der Uniklinik Köln
Department of General, Visceral, Cancer, and Transplantation Surgery
Kerpener Str. 62, 50937 Köln, Germany
PD Florian Gebauer, MD

*Rostock:*
**Klinikum Südstadt Rostock**
Department of Surgery
Südring 81, 18059 Rostock, Germany
Prof Kaja Ludwig, MD

*Berlin:*
**Helios Klinikum Emil von Behring**
Department of Surgery
Walterhöferstraße 11, 14165 Berlin, Germany
Prof Marc H. Jansen, MD

*Kaiserslautern:*
**Westpfalz-Klinikum GmbH**
Department of General, Visceral and Transplantation Surgery
Hellmut-Hartert-Straße 1, 67655 Kaiserslautern, Germany
Stefan Bergheim, MD

*Westerstede:*
**Ammerland-Klinik GmbH**
Klinik für Allgemein- und Viszeralchirurgie
Lange Str. 38, 26655 Westerstede, Germany
Muneer Deeb, MD

*Bochum:*
**St. Josefs-Hospital**
Department of General and Visceral Surgery, Ruhr-University Bochum
Gudrunstraße 56, 44791 Bochum, Germany
Monika Janot-Matuschek, MD

*Marburg:*
**University Hospital Marburg**
Department of Visceral, Thoracic- and Vascular Surgery, Philipps-University Marburg
Baldingerstraße 1, 35043 Marburg, Germany
Prof Detlef K. Bartsch, MD

*Bremen:*
**Klinikum Bremen-Mitte**
Department of General, Visceral and Oncological Surgery, Hospital Group Gesundheit Nord
St. Jürgen-Straße 1, 28205 Bremen, Germany
Prof Hüseyin Bektas, MD

*Osnabrück:*
**Niels-Stensen-Kliniken Marienhospital Osnabrück**
Department of General and Visceral Surgery
Bischofsstr. 1, 49074 Osnabrück, Germany
Prof Christoph Nies, MD

*Lübeck:*
**University Medical Center Schleswig-Holstein, Campus Luebeck**
Department of Surgery,
Ratzeburger Allee 160, 23538 Lübeck, Germany
Prof Tobias Keck, MD

*Schwerin:*
**Helios Klinik Schwerin**
Department of Surgery
Wismarsche Str. 393–397, 19049 Schwerin, Germany
Franziska Koch, MD

*Regenburg:*
**Krankenhaus Barmherzige Brüder**
Department of General and Visceral Surgery
Prüfeninger Str. 86, 93049 Regensburg, Germany
Prof Pompiliu Piso, MD

*Munich:*
**München Klinik Neuperlach**
Department of Surgery
Oskar-Maria-Graf Ring 51, 81737 Munich, Germany
Prof Natascha Nüssler, MD

*Villingen-Schwenningen*
**Schwarzwald-Baar-Klinikum**
Department of Surgery
Klinikstr. 11, 78052 Villingen-Schwenningen
Prof Stefan Beckert, MD

***----------- Not recruiting yet ------------------***

***Dortmund:***
**Klinikum Dortmund**
Department of Surgery
Beurhausstrasse 40, 44137 Dortmund, Germany
Prof Maximilian Schmeding, MD MHBA

***Düsseldorf:***
**Evangelisches Krankenhaus Düsseldorf**
Department of Surgery
Kirchfeldstr. 40, 40217 Düsseldorf, Germany
Prof Werner Hartwig, MD

***Nuremberg:***
**Paracelsus Medical University Nuremberg**
Department of Surgery
Professor-Ernst-Nathan-Str. 1, 90419 Nürnberg, Germany
Marcus Renz, MD

***Frankfurt:***
**University Hospital Frankfurt, Goethe University Frankfurt**
Department of General, Visceral and Transplant Surgery
Theodor-Stern-Kai 7, 60590 Frankfurt, Germany
Prof Otto Wolf Bechstein, MD

***Bonn:***
**University Hospital Bonn**
Department of Surgery
Venzsberg-Campus 1, 53127 Bonn
Prof Steffen Manekeller, MD

***Stuttgart:***
**Stuttgart Clinics – Katharinenhospital**
Department of General, Visceral, Thoracic and Transplantation Surgery
Kriegsbergstraße 60
70174 Stuttgart
Prof Jörg Köninger, MD

□
