## Supplementary material for "Pylorus resection versus pylorus preservation in pancreatoduodenectomy (PyloResPes): study protocol and statistical analysis plan for a German multicentre, single-blind, surgical, registry-based randomised controlled trial": PyloResPres__SupplementalFile_2_StuDoQ Registry

### SUPPLEMENTARY MATERIALS

#### StuDoQ|Pancreas registry:
#### Further information regarding data collection

The StuDoQ software environment is used for documenting routinely collected clinical data for the StuDoQ|Pancreas registry. Participating hospitals receive a comprehensive statistical evaluation of their data, including a comparison with the (anonymous) overall registry data. Thereby, a valid and up-to-date "benchmarking" is possible.

A very flexible adjustable error and plausibility checking ensures data quality. Entered clinical data can be exported and downloaded by the participating institution at any time. Various export formats (Excel; SPSS; CSV) are available. Due to these technical export opportunities, a high level of transparency is guaranteed, as an analysis of the entered data at the institutional level is possible at any time, without special request to the DGAV or any external services. Training courses regarding documentation and evaluation for staff of all participating hospitals are offered by the DGAV two or three times per year. Additional information and guidance to trial-related questions will be provided to participating centres and affiliated staff in a timely manner.

The StuDoQ registry provides the technical infrastructure to optimally support data collection: data entry is embedded within a convenient user-friendly website. Additional information (help texts, current news, etc.) can be displayed, which can be easily added or modified within the content management system at any time. In addition, participating institutions will be continuously notified by email regarding e.g., eCRF changes.[1]

1. DGAV. StuDoQ – German Society of General and Visceral Surgery [Deutsche Gesellschaft für Allgemein- und Viszeralchirurgie e.V. (DGAV)] [Available from: http://www.dgav.de/studoq].
