## Supplementary material for "Pylorus resection versus pylorus preservation in pancreatoduodenectomy (PyloResPes): study protocol and statistical analysis plan for a German multicentre, single-blind, surgical, registry-based randomised controlled trial": PyloResPres__SupplementalFile_3_StatisticalMethods

### SUPPLEMENTARY MATERIALS

### Statistical and methodological considerations

#### 1 Definition of primary outcome: DGE occurrence

The primary outcome is DGE graded according to the International Study Group of Pancreatic Surgery (ISGPS) consensus definition. Grade A is defined as either nasogastric tube (NGT) insertion after POD 3 or as the inability to tolerate solid diet intake by POD 7. Grade B is defined as using a NGT for 8–14 days, NGT reinsertion after POD 7, or the inability to tolerate a solid diet by POD 14. Grade C is defined as the need to use an NGT for more than 14 days, NGT reinsertion after POD 14, or the inability to tolerate a solid diet by POD 21. DGE of grades B and C are considered to be clinically relevant DGE grades.

However, in this trial the overall DGE occurrence within the assessment period of 30 postoperative days will be compared between both intervention groups. Hence, patients who develop a DGE will be classified as DGE positive irrespective of the grading.

#### 2 Pre-trial sample size calculation – further details

The effectiveness of the two competing PD techniques (pylorus-resecting (prPD) vs pylorus-preserving (ppPD)) against DGE will compared. Due to relevant uncertainties of the rates regarding the primary endpoint "*occurrence of DGE up to POD 30 (all grades)*" reported in the current literature, a two-stage group sequential design was set up, with one interim analysis after assessment of 50% of the initial target sample size, thus implying a multiple test problem. The critical values and the test characteristics of this group sequential test design were calculated for the optimum design within the Wang & Tsiatis class, with boundary shape parameter Delta = 0.23 (values of Delta between 0 and 0.5 give critical values ranging between Pocock and O'Brien & Fleming tests).[1,2] This Delta minimizes the optimization criterion $ASN_{H0}$ + $ASN_{H01}$ + $ASN_{H1}$, whereas $ASN_{H0}$ and $ASN_{H1}$ is the expected sample size or average sample number (ASN) under the null and alternative hypothesis, respectively; $ASN_{H01}$ denotes the expected sample size calculated midway between H0 and H1, i.e., for Delta/2.

For a pre-specified significance level of 5% (two-sided) and event rates $\pi_{prPD}$ =0.3 and $\pi_{ppPD}$ =0.4 (odds ratio (OR) of 1.556), the power is 90.0% if the test stages consist of the sample sizes given in the last two columns of **table S1** (Fisher's exact test applied). Given the planned allocation ratio of 1:1, this yields a total of 984 patients (2×492) to be allocated to this superiority trial. For comparison, the maximum sample size in the group sequential test design is 1.03 times the sample size in a corresponding fixed sample size design (2×476 patients). The interim analysis will be performed at the 50% information fraction, i.e. after **492 patients** in total (246 patients randomized per group) (Software used: ADDPLAN® V6.1.1). A total number of **984 patients** will be assigned to the trial, taking into account 15% missing information with respect to DGE due owing to intraoperative discovery of an extended or unresectable disease or due to loss to follow-up after PD (expected to be extremely low). Assuming that about 70% of eligible patients consent to study participation, we expect to screen about 1404 patients in total (**Figure 2**).

**Table S1.** Sequential analysis with a maximum of 2 looks (group sequential design), i.e. 1 interim stage and a final stage**.** Boundary values according to the Wang & Tsiatis power family.

| Information rate | bounds accept $H_0$ | bounds reject $H_0$ | sign.level one-sided | α spent | β spent | power achieved | stage $n_1$ | sizes $n_2$ |
|---|---|---|---|---|---|---|---|---|
| 0.5 | - | 2.449 | 0.0072 | 0.0143 | - | 0.4516 | 245.3 | 245.3 |
| 1.0 | 2.031 | 2.031 | 0.0211 | 0.0500 | - | 0.9000 | 245.3 | 245.3 |

Bounds indicate critical values.

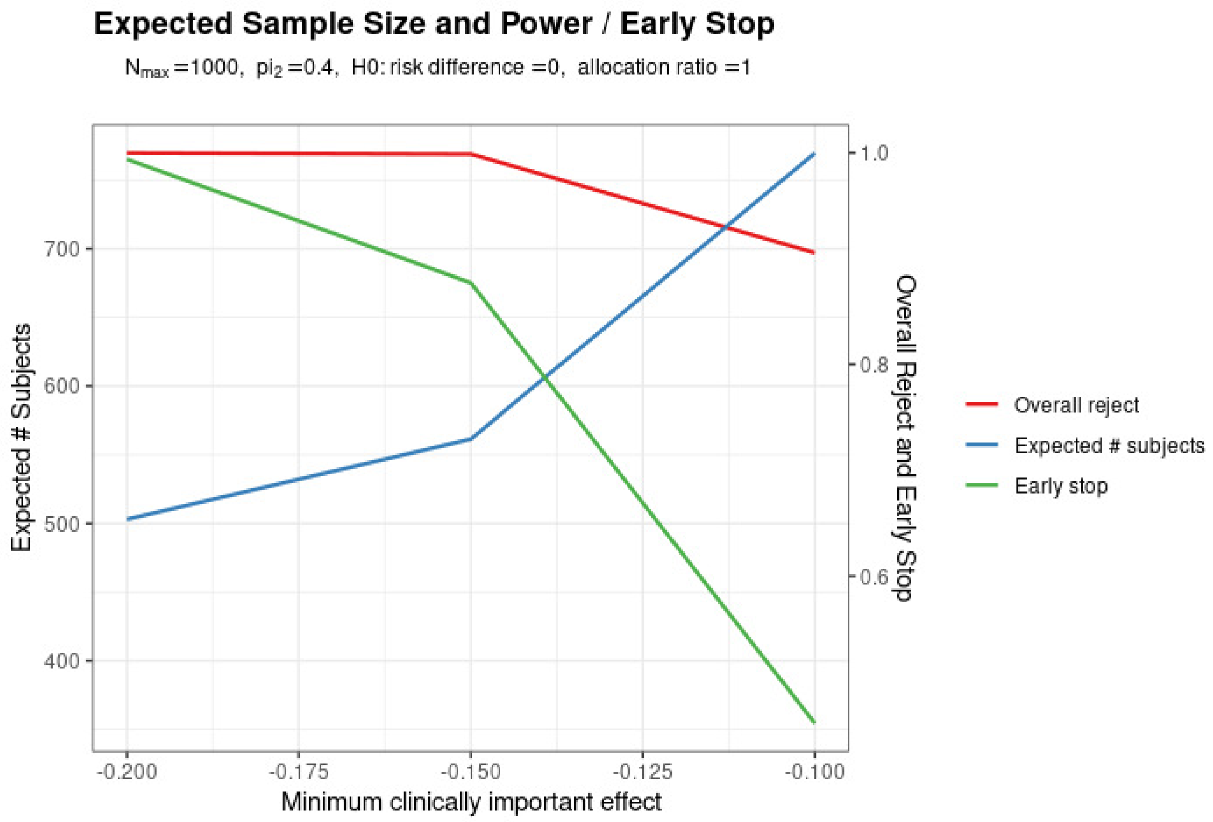


**Figure 1. Plot of average expected sample size and power for a Wang & Tsiatis design with two looks**.[3] Minimum clinically important effect of 0.2, 0.15, or 0.1, derived for a DGE rate $pi_1$ = {0.2, 0.25, 0.3} in the experimental group (prPD). $pi_2$ = 0.4 depicts the DGE rate in the reference group (ppPD).
ppPD = Pylorus preservation in pancreatoduodenectomy; prPD = Pylorus resection in pancreatoduodenectomy.

□
